# The Epidemiological and Economic Burden of Diabetes in Ghana: A Scoping Review to Inform Health Technology Assessment

**DOI:** 10.1101/2023.04.19.23288806

**Authors:** Mohamed Gad, Joseph Kazibwe, Emmanuella Abassah-Konadu, Ivy Amankwah, Richmond Owusu, Godwin Gulbi, Sergio Torres-Rueda, Brian Asare, Anna Vassall, Francis Ruiz

## Abstract

**Introduction:** Diabetes remains one of the four major causes of morbidity and mortality globally among non-communicable diseases (NCDs. It is predicted to increase in sub–Saharan Africa by over 50% by 2045. The aim of this study is to identify, map and estimate the burden of diabetes in Ghana, which is essential for optimising NCD country policy and understanding existing knowledge gaps to guide future research in this area.

**Methods:** We followed the Arksey and O’Malley framework for scoping reviews. We searched electronic databases including Medline, Embase, Web of Science, Scopus, Cochrane and African Index Medicus following a systematic search strategy. The Preferred Reporting Items for Systematic Reviews and Meta-Analyses Extension for Scoping Reviews was followed when reporting the results.

**Results:** A total of 36 studies were found to fulfil the inclusion criteria. The reported prevalence of diabetes at national level in Ghana ranged between 2.80% – 3.95%. At the regional level, the Western region reported the highest prevalence of diabetes: 39.80%, followed by Ashanti region (25.20%) and Central region at 24.60%. The prevalence of diabetes was generally higher in women in comparison to men. Urban areas were found to have a higher prevalence of diabetes than rural areas. The mean annual financial cost of managing one diabetic case at the outpatient clinic was estimated at GHS 540.35 (2021 US $194.09). There was a paucity of evidence on the overall economic burden and the regional prevalence burden.

**Conclusion:** Ghana is faced with a considerable burden of diabetes which varies by region and setting (urban/rural). There is an urgent need for effective and efficient interventions to prevent the anticipated elevation in burden of disease through the utilisation of existing evidence and proven priority-setting tools like Health Technology Assessment (HTA).

## Introduction

Diabetes is one of the top four non-communicable diseases (NCDs) in terms of mortality globally^1^. Approximately 537 million people between the ages of 20-79 years are living with diabetes globally of which over 75% live in low- and middle-income countries (LMICs). Of those living with diabetes nearly half are unaware of their diagnosis^2^. Diabetes exerts tremendous pressure on the resources available for health; treatment and management of diabetes account for over 10% of the total health expenditure among adults globally^3^. The prevalence of diabetes is expected to increase globally to 783 million by 2045^2^.

As of 2021, there were approximately 24 million people with diabetes in sub-Saharan Africa (SSA) and the number is projected to increase by 134% by 2045. ^3^. The prevalence of diabetes in the region stands at approximately 4.5% among those aged between 20-79 years. In 2021 alone, over 306,000 people under 60-years of age died due to diabetes in SSA^3^. It is estimated that each person with diabetes incurs approximately USD 547 per year on healthcare (both patient and health system direct costs) in SSA as of 2021.

Most cases of diabetes can be classified into two types: Type 1 diabetes (T1D) is most common in children and results from the destruction of insulin-producing beta cells, mostly by autoimmune mechanisms^4^. Type 2 diabetes (T2D) is a metabolic disorder characterised by insulin resistance and relative insulin deficiency and, although it can occur at any age, it is most common among adults^5^. T2D is linked to physical inactivity and an unhealthy diet, and accounts for approximately 90% of all diabetes cases globally^6^.

While regional estimates are available to support policy making, there is a need to understand the country-specific disease burdens (national and sub-national) to inform target policy at a local level that yields effective and economically efficient impact. Unfortunately, existing estimates of country-specific diabetes burden are sparse in low and lower middle-income settings, especially in SSA. This makes it difficult to identity and implement appropriate targeted interventions that are feasible and affordable by a given country considering the existing financial constraints of the health budgets. Countries in SSA are starting to adopt health technology assessment (HTA) as a decision-making aid to identify and implement appropriate interventions while maximizing value for money. Figure 1 illustrates the main steps involved in HTA processes as adapted from Siegfried et al.7

**Figure 1:**
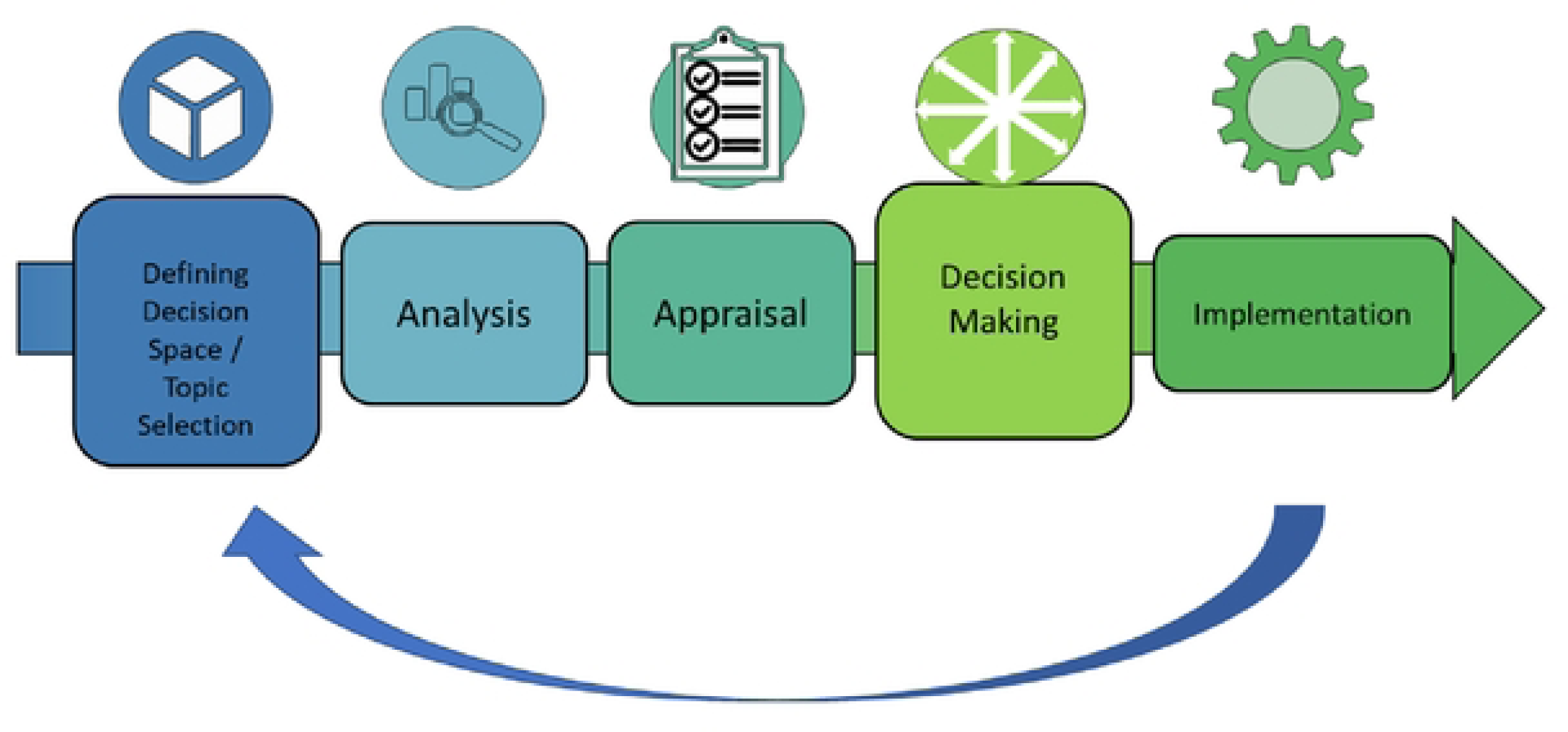
The HTA process.

HTA can be defined as a multidisciplinary process that uses explicit methods, often involving cost-effectiveness analysis, to determine the value of a health technology at different points in its lifecycle^8^. The HTA process typically starts with identifying relevant alternatives linked to a policy need (defining the decision space). This is then followed by the gathering and synthesis of various sources of evidence to arrive at an understanding of the relative value for money of the alternatives (the analysis or assessment step). There is then an appraisal of that evidence involving some form of deliberative process against decision criteria, leading to the development of recommendations and the subsequent implementation of preferred options. Information gathered from implementation can be used to inform a future HTA. An important input into the conduct of HTA includes having credible estimates of disease burden, which will be important in informing key parameters in any cost-effectiveness model^9^.

Ghana has adopted the use of HTA to inform decision making within the health sector^10^. This commitment to evidence-informed priority setting has included the development and implementation of an HTA process guide, officially launched in December 2022^11^. The Ghana Ministry of Health has indicated an interest in using HTA to inform decision making in diabetes management and prevention. Such an approach requires up-to date information about the current epidemiological and economic burden of diabetes in the country, which is currently unavailable^12^.

To address this need, our study aims to undertake a scoping review of the peer-reviewed literature to identify, map and estimate the burden of diabetes in Ghana, in terms of epidemiological distribution, health outcomes and economic consequences. It is anticipated this work will support optimising current NCD country policy, especially in relation to priority setting, as well inform the parameterisation of model-based analyses and highlight existing knowledge gaps to guide future research in this area.

## Methods

### Study design

We followed the Arksey and O’Malley framework for scoping reviews^13^. The framework consists of five stages which were followed: (i) identifying the research question; (ii) identifying relevant studies; (iii) selecting appropriate studies; (iv) charting and collating the data, and (v) summarising and reporting the results. The detailed search protocol is available in Appendix 1. A study protocol is available although not registered. The Preferred Reporting Items for Systematic Reviews and Meta-Analyses Extension for Scoping Reviews (PRISMA-ScR) was followed when reporting the results^14^. We examined the following aspects of burden: (1) epidemiological distribution (incidence, prevalence, demographic distribution), (2) impact on health outcomes (comorbidity or complications, health effects in disability adjusted life years (DALYs), quality adjusted life years (QALYs), mortality among others), and (3) economic consequences (cost of care, loss of productivity, or out of pocket expenditure).

### Eligibility criteria and study selection

A study was considered eligible for inclusion if it was published in a peer-reviewed journal and reported burden of disease in Ghana reflecting at least one of the three dimensions highlighted above. To ensure relevance, we only included studies published after 2009. All study designs were considered for inclusion without restrictions. There were also no restrictions on population, age or sex. Literature reporting only the qualitative experience of diabetic patients, and those assessing the relationship between socioeconomic status (SES), gender, and diabetes as a health outcome were excluded. Studies published in languages other than English were also excluded.

### Information Sources

We searched the following electronic databases: Medline, Embase, Web of Science, Scopus, Cochrane and African Index Medicus. The databases were searched on 4^th^ April 2021 following a systematic search strategy and a second search was done on 11^th^ April 2023 to find all new articles that were published since the previous search.

### Search strategy

We used two broad key terms (‘Ghana’ and ‘diabetes’) as well as similar derivatives, to identify the literature on the burden of disease. Search strings were tailored to the different databases (appendix 1).

### Selection process

The retrieved articles from the search were listed and uploaded to Covidence software^15^ which was used to identify and remove duplicates, carry out the screening process and full-text review. A standard process of screening articles by title and abstract, followed by full-text reading was followed to assess eligibility to be included in this study. These steps were conducted by two independent researchers (MG & JK). Any discrepancies in the assessment decision were discussed and resolved by reaching a consensus between the two researchers.

### Data charting

A data extraction sheet was used to extract relevant information from included studies to allow us to map and highlight the main results and categorize findings in relation to the research question. Information extracted included: author name, publication year, form of diabetes burden being reported (such as prevalence, incidence, economic, etc), study design, target population, geographical region, setting (urban or rural), and main findings.

Our charting approach allowed us to interpret data from included studies according to the forms of diabetes burden which we henceforth refer to as themes. The extracted data was grouped under three themes (epidemiological, health outcomes, economic). We used simple visualisation and basic descriptive analytical techniques to summarise and report the scoping review findings.

### Synthesis of results

We organised extracted data quantitatively following the themes. We produced tables and charts in relation to the following: the distribution of studies geographically and per type of burden, target groups; the research methods adopted, and health outcome measures used.

## Results

The electronic database search yielded 1103 records after deduplication. Figure 2 below shows the PRISMA flowchart.

**Figure 2:**
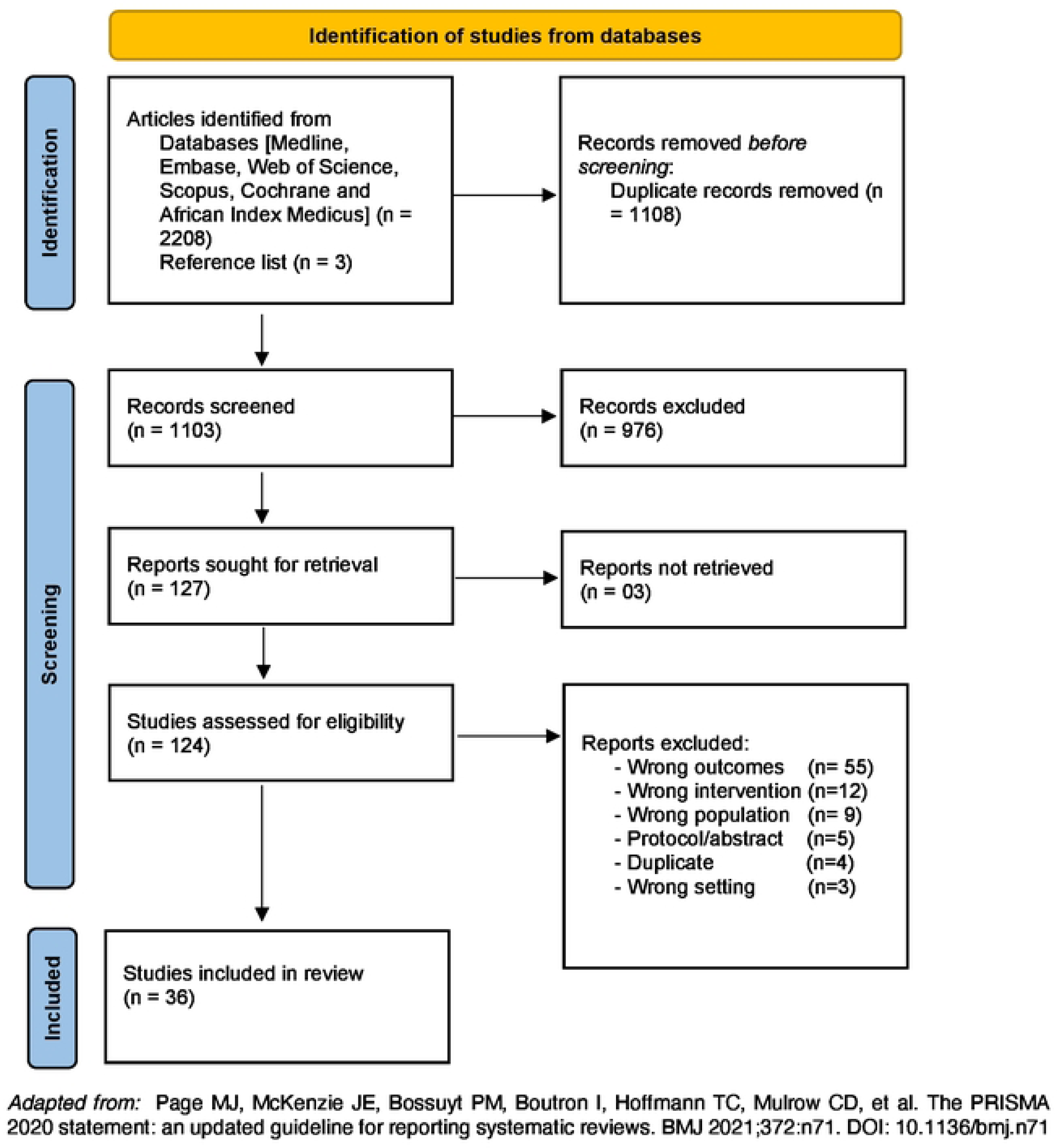
PRISMA flow chart.

### Study characteristics

A total of 36 studies fulfilled the inclusion criteria. Table 1 shows the characteristics of the included studies. All studies were observational studies with a majority having a cross-sectional design (n=30); there were four longitudinal studies^16–19^ and two case control studies^20,21^. The longitudinal studies were based on either panel data or cohort study data.

**Table 1:** Table of study characteristics.

| Author | Publication year | Study design | Target population | Form of the Burden | Specific burden | Region |
| --- | --- | --- | --- | --- | --- | --- |
| Gatimu et al.<sup>22</sup> | 2016 | Cross-sectional | Adults 50 years and above | Prevalence | Prevalence of diabetes and risk factors | National |
| Bawah et al.<sup>26</sup> | 2019 | Cross-sectional | Adults 30 years and above | Prevalence | Prevalence of T2D and pre-diabetes | Volta region |
| Kojo Anderson<sup>38</sup> | 2017 | Cross-sectional | Adults 18 years and above | Prevalence | Prevalence of T2D | Western Region, Central Region |
| Vuvor et al.<sup>30</sup> | 2011 | Cross-sectional | Adults 36 years and above | Prevalence | Prevalence of diabetes and risk factors | Greater Accra |
| Chilunga et al.<sup>39</sup> | 2019 | Cross sectional | Adults 25 -70 years with BMI<25kg/m2 | Prevalence | Prevalence of T2D | Ashanti region |
| Gato et al.<sup>33</sup> | 2017 | Cross-sectional | Adults 18 years and above | Prevalence | Prevalence of diabetes | Central Region |
| Yorke et al.<sup>23</sup> | 2020 | Cross-sectional | Adults 50 years and above | Prevalence | Prevalence of diabetes | National |
| Agyemang et al.<sup>40</sup> | 2016 | Cross sectional | Adults 25 -70 years | Prevalence | Prevalence of T2D and Obesity | Ashanti region |
| Tyrovolas et al.<sup>36</sup> | 2015 | Cross-sectional | Adults 18 years and above | Prevalence | Prevalence of diabetes | National |
| Annani-Akollor et al.<sup>41</sup> | 2019 | Cross-sectional | Adults 18 years and above | Complications | Complications of T2D: Macrovascular, microvascular, neuropathy, nephropathy, retinopathy, sexual dysfunction, DKA, hypoglycemia | Ashanti region |
| Hayfron-Benjamin et al.<sup>37</sup> | 2019 | Cross-sectional | Adults above 25 years with T2D | Complications | Complications of T2D: Macrovascular, microvascular, coronary artery disease, nephropathy, retinopathy, PAD, stroke | National |
| Nsiah et al.<sup>42</sup> | 2015 | Cross-sectional | Adults 20 - 80 years with T2D | Complications | Prevalence of comorbidities and risk factors (Metabolic Syndrome, hypertension, dyslipidemia) | Ashanti region |
| Mogre et al.<sup>34</sup> | 2014 | Cross-sectional | Adults 18 years and above with T2D | Comorbidities | Prevalence of MetS among diabetic patients | Northern region |
| Agyemang-Yeboah et al<sup>43</sup> | 2019 | Cross-sectional | Patients with diabetes | Comorbidities | Prevalence of MetS among diabetic patients | Ashanti Region |
| Antwi-Bafour et al<sup>20</sup> | 2016 | case-control | Patients with T2D | Comorbidities | Prevalence of anaemia among diabetics | Not mentioned |
| Akpalu et al<sup>31</sup> | 2018 | Cross-sectional | Adults 30 -65 years with T2D | Comorbidities | Prevalence of depression among T2D patients | Greater Accra |
| Sarfo et al<sup>16</sup> | 2018 | Longitudinal (Cohort) | Adults 18 years and above | Comorbidities | Prevalence of stroke among diabetics | Eastern, Ashanti, Northern Regions |
| Osei-Yeboah et al<sup>27</sup> | 2017 | Cross-sectional | Patients with T2D | Comorbidities | Prevalence of MetS among diabetic patients | Volta region |
| Opere-Addo et al<sup>44</sup> | 2020 | Cross-sectional | Adults 18 years and above | Comorbidities | Prevalence of hypertension among diabetics | Ashanti region |
| Sarfo-Kantanka et al<sup>17</sup> | 2016 | Longitudinal (panel) | Patients with T2D admitted to hospital | Mortality | Mortality trend for 31 years | Ashanti region |
| Lartey & Aikins<sup>45</sup> | 2018 | Cross-sectional | Patients with diabetes attending Diabetic clinic | Comorbidities | Prevalence of visual impairment among diabetics | Ashanti region |
| Nimako et al<sup>32</sup> | 2013 | Cross-sectional | Patients with T2D attending General Hospital | Prevalence and comorbidities | Prevalence of diabetes and hypertension | Greater Accra Region |
| Atosona & Larbie<sup>46</sup> | 2019 | Cross-sectional | Patients with T2D at the outpatient clinic | Complications | Prevalence of diabetic foot | Greater Accra, Ashanti, and Northern regions |
| Sarfo-Kantanka et al<sup>18</sup> | 2019 | Longitudinal (Cohort) | Patients with T2D at the diabetes clinic | Incidence | Incidence rate of diabetes-related LLA | Ashanti region |
| Sarfo-Kantanka et al<sup>19</sup> | 2018 | Longitudinal (panel) | Patients with T2D at the diabetes clinic | Incidence | Trend of incidence and predictors of diabetic foot | Ashanti region |
| Quaye et al<sup>25</sup> | 2015 | Cross-sectional | Patients with diabetes | Economic | Annual costs | Greater Accra, Ashanti, Eastern Regions |
| Tarekegne et al<sup>24</sup> | 2018 | Cross-sectional | Adults 50 years or above | Prevalence | Prevalence of DM | National |
| Cook-Huynh et al<sup>47</sup> | 2012 | Cross-sectional | Adults 18 years or above | Prevalence | Prevalence of DM | Ashanti region |
| Sarfo-Kantanka et al<sup>48</sup> | 2014 | Cross-sectional | Adults 18 years or above | Prevalence | Prevalence of DM | Ashanti region |
| Agbogli et al<sup>49</sup> | 2017 | Cross-sectional | Adults 18 years or above | Prevalence | Prevalence of DM | Ashanti region |
| Quaicoe et al<sup>21</sup> | 2017 | case-control | Adults aged 18-64+ years | Prevalence | Prevalence of DM | Ashanti region |
| Odame Anto et al<sup>50</sup> | 2021 | Cross-sectional | Adults aged 30 years or above | Complications | Prevalence of MetS among diabetic patients | Ashanti region |
| Abagre et al<sup>35</sup> | 2022 | Cross-sectional | Adults aged 30-79 years old | Complications | Prevalence of MetS among diabetic patients | Brong-Ahafo region |
| Abu et al<sup>51</sup> | 2022 | Cross-sectional | Adults aged 38-85 years old | Complications | Prevalence of dry eye disease among T2D patients | Central region |
| Tuglo et al<sup>28</sup> | 2022 | Cross-sectional | Adults | Complications | Prevalence of diabetic ulcers | Volta region |
| Ellahi et al<sup>29</sup> | 2022 | Cross-sectional | Adults 18 years or above | Prevalence | Prevalence of DM | Volta region |

All studies included adults (persons aged 18 and above) as their target population. Three studies focused on people above the age of 50 years^22–24^. Forms of burden reported in the studies were prevalence (n=16), complications and comorbidity (n=17), incidence (n=2)^18,19^, economic (n=1)^25^ and mortality (n=1)^17^. All studies either reported on T2D or diabetes in general (without specifying the type).

Notably, most of the extracted studies were carried out in Ashanti region (n=14), followed by Volta (n=4)^26–29^, Greater Accra (n=3)^30–32^, Central (n=1)^33^, Northern (n=1)^34^ and Brong Ahafo (n=1)^35^. Four studies were carried out in more than one region while five studies took a whole country perspective^22–24,36,37^. No study was carried out that specifically focused on the following regions: Upper East, Upper West, Western, and Eastern.

### Publication trend

The number of publications by year (figure 3) broadly shows an increasing trend between 2009 and 2019 and a dip after 2019. Most articles were published in 2019 (n=7) before a decline in 2020.

**Figure 3:**
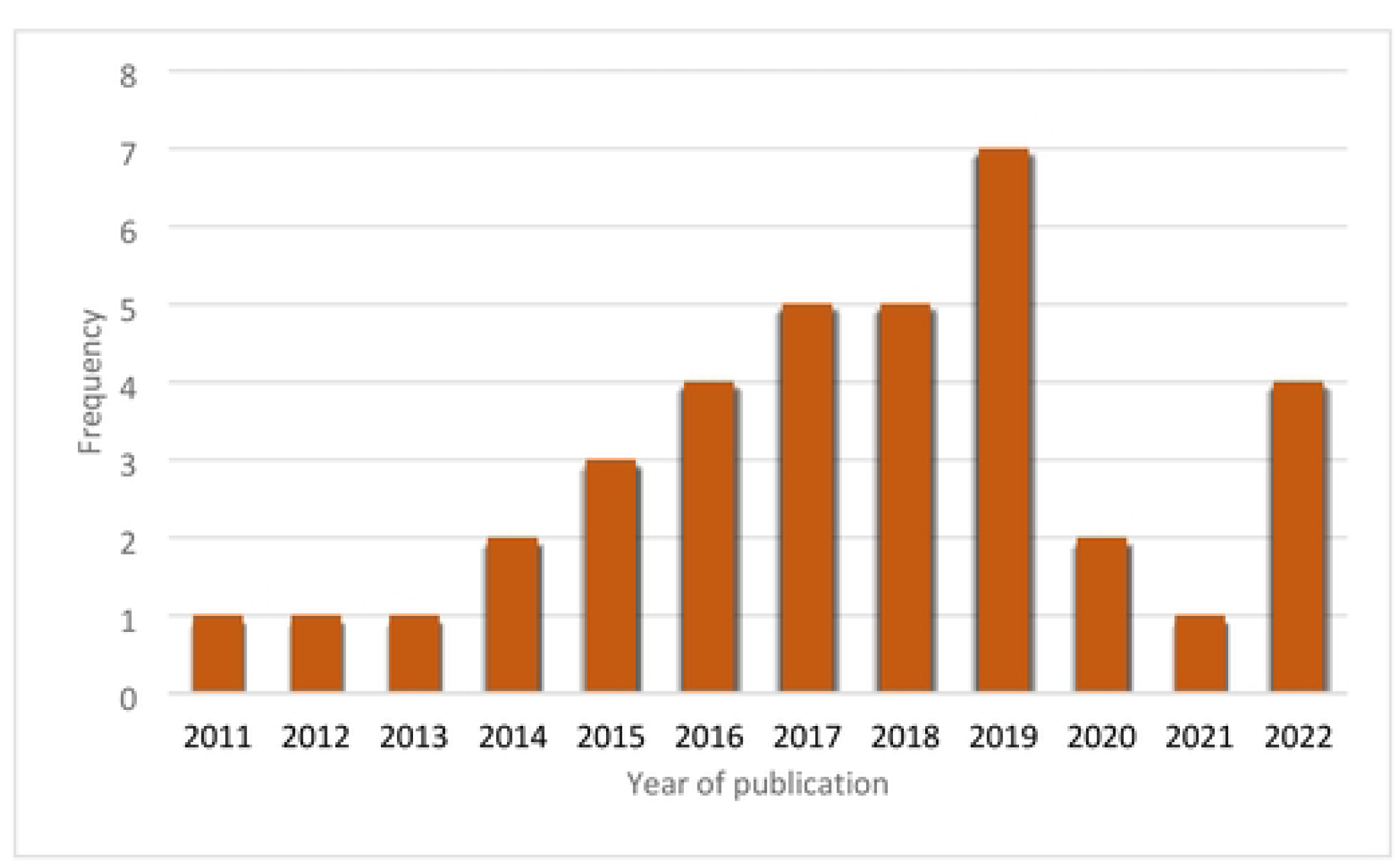
Number of studies/publications per year.

### Forms of the burden of diabetes (themes) in Ghana

Our charting analysis mapped the studies across three main themes: epidemiological, health outcomes, and economic. The epidemiological theme included prevalence of diabetes (n=16) and incidence (n=2). The health outcomes theme (n=17) was comprised of studies reporting on complications (n=6), comorbidities (n=10) and mortality (n=1). Finally, the economic theme (n=1) included costs of diabetes services. Some studies reported both the prevalence of diabetes and comorbidities (n=1). Figure 4 demonstrates the percentage of included studies by theme.

**Figure 4:**
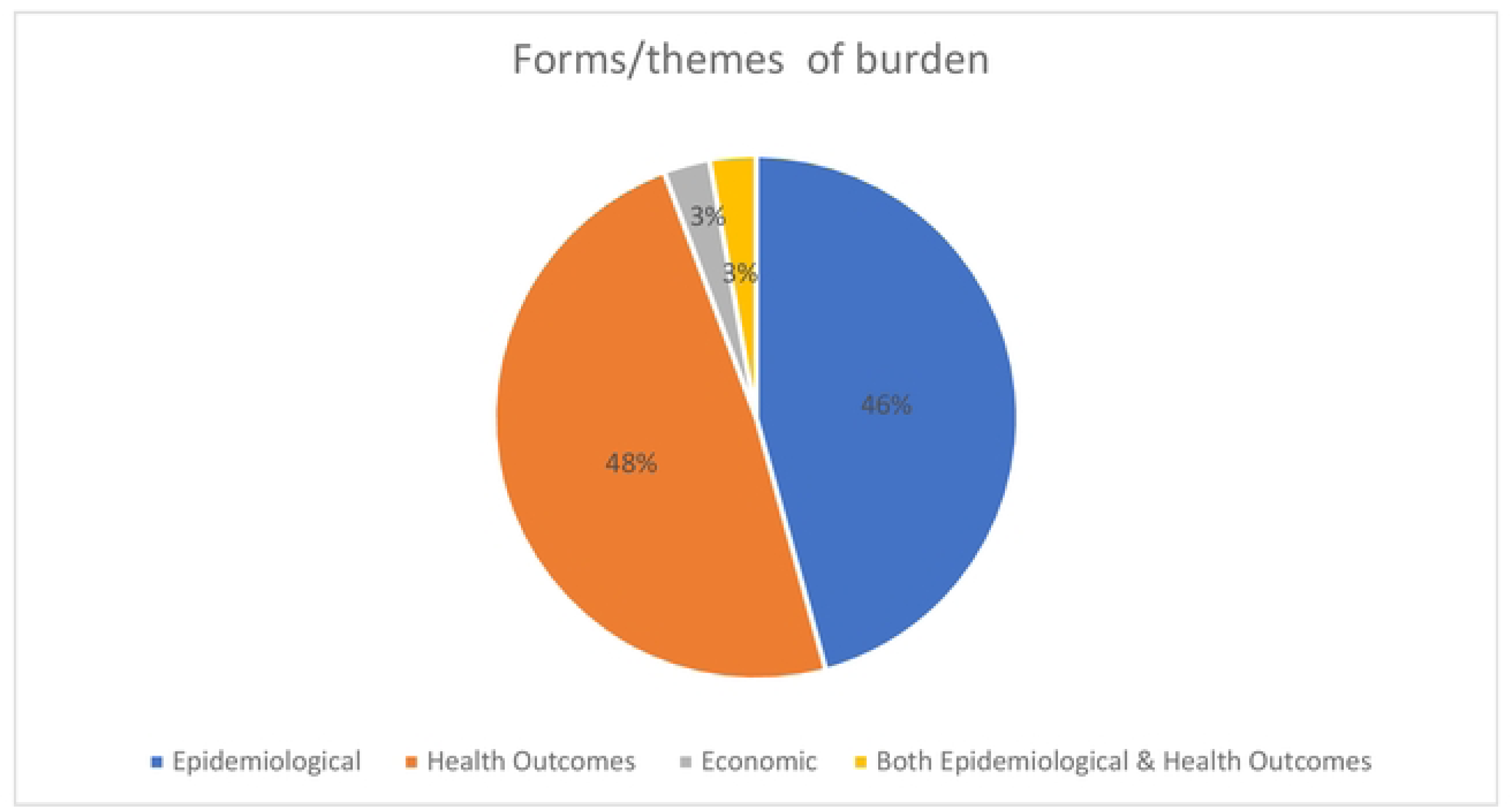
Percentage of included studies by form of burden (theme)

### Frequency of theme by region

The Ashanti and Greater Accra regions were the most frequently targeted regions across all studies, with reported studies covering all the three themes. We found data on both prevalence and outcomes in the Volta and Central regions. The Western region had only prevalence data reported while Brong-Ahafo and Northern regions had studies that reported on health outcomes only. No data was found for the remaining regions. Five studies were conducted using a nationally representative sample. Figure 5 shows the frequency of themes by region from all included studies (including single and multi-region studies).

**Figure 5:**
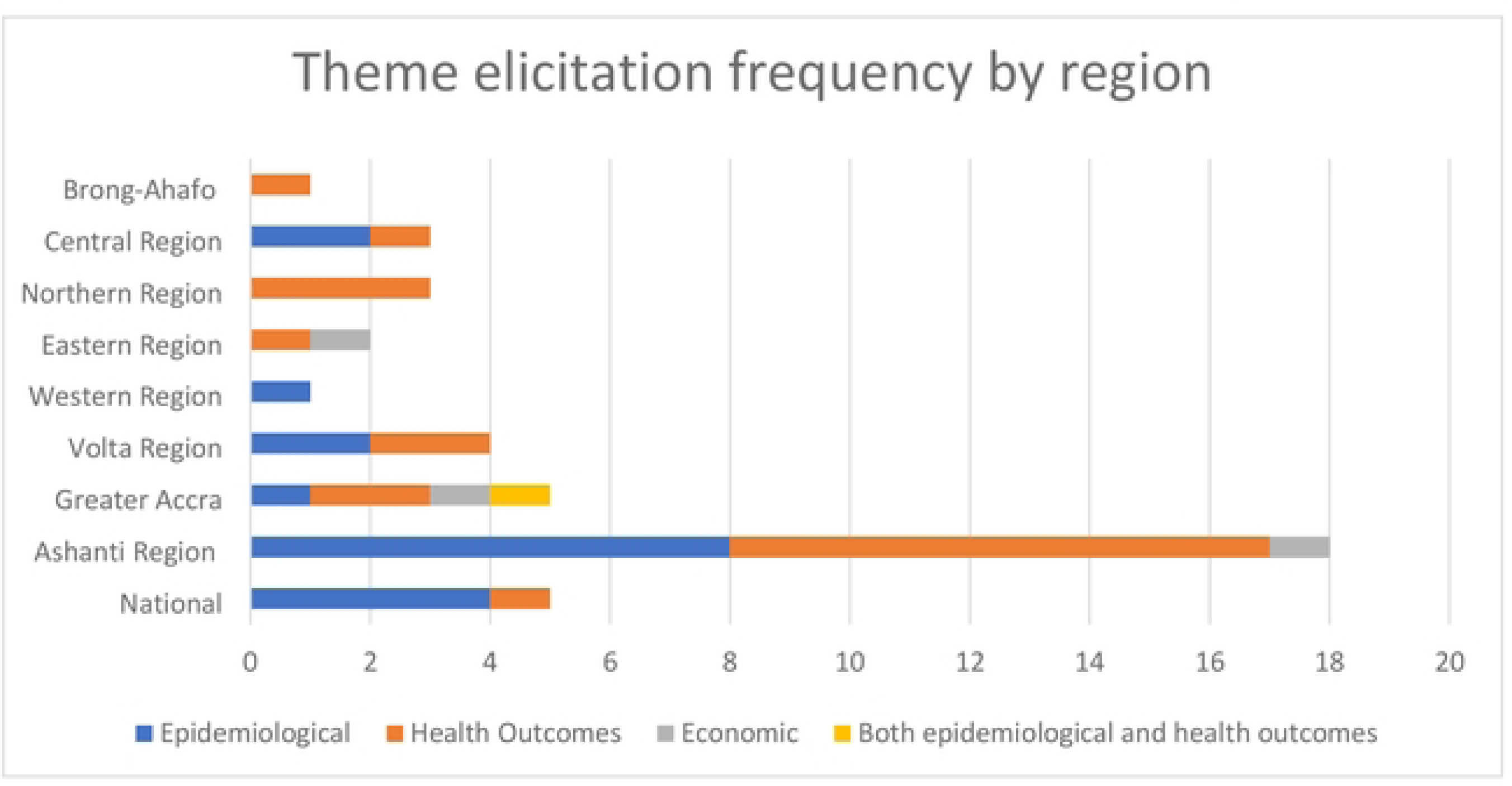
Frequency of burden of disease themes reported by region.

#### 1) Epidemiological burden

The reported prevalence of diabetes at national level in Ghana ranged between 2.80%^23,24^ – 3.95%^22^. At sub-national levels, the Western region reported the highest prevalence of diabetes: 39.80% among those 18 years and older^38^. The second highest prevalence of diabetes (25.20%) was reported in Ashanti region^48^ followed by 24.60% in the Central region^38^.

In the national studies, the prevalence of diabetes was generally higher in women in comparison to men^22,24,36^. Regionally, the prevalence of diabetes was also notably higher in females compared to males, with the exception of the Ashanti region^26,30,48,49^. Urban areas had a higher prevalence than rural areas^22–24,39,40^.

Prevalence studies only covered five of the ten administrative regions: Ashanti (n=6), Volta (n=3), Central (n=2), Greater Accra (n=2), and Western (n=1) regions. There were four national-level studies. The majority of prevalence studies were focused on adults aged 18 years old and above (n=7), followed by adults 50 years and above (n=4). Table 2 shows the summary of studies that reported the prevalence of diabetes in Ghana.

**Table 2:** Summary of findings of studies reporting the prevalence of diabetes in Ghana.

| Author and Year | Region | Age groups | Sample size | Diagnostic criteria | Prevalence % (95% CI) |  |  |  |  |
| --- | --- | --- | --- | --- | --- | --- | --- | --- | --- |
|  |  |  |  |  | Overall | Male | Female | Urban | Rural |
| Tyrovolas et al. 2015<sup>36</sup> | National | Adults 50 years or above | 52,946 | Self-reported | 3.9 (2.4 - 6.2) | NR | NR | NR | NR |
| Gatimu et al. 2016<sup>22</sup> | National | Adults 50 years or above | 4,089 | Self-reported | 4.0 (3.4 - 4.6) | 1.7 (1.3–2.3) | 2.2 (1.7 - 2.8) | 6.2 (4.8 - 8.0) | 2.3 (1.7-3.3) |
| Tarekegne et al. 2018 | National | Adults 50 years or above | 4,289 | Self-reported | 2.8 | 2.5 | 3.3 | 4.7 | 1.5 |
| Yorke et al. 2020<sup>23</sup> | National | Adults 50 years or above | 3,350 | Self-reported | 2.8 (2.0 - 3.9) | 2.8 | 2.8 | 4.4 | 1.3 |
| Cook-Huynh et al. 2012<sup>47</sup> | Ashanti | Adults 18 years or above | 326 | WHO diagnostic criteria | NR | NR | NR | NR | 7.7 (5.0-11.0) |
| Sarfo-Kantanka et al. 2014<sup>48</sup> | Ashanti | Adults 18 years or above | 1,292 | WHO diagnostic criteria (fasting blood glucose only) | 25.2 | 25.7 | 24.4 | NR | NR |
| Agyemang et al. 2016<sup>40</sup> | Ashanti | Adults aged 25-70 years old | 820 | WHO diagnostic criteria | NR | NR | NR | 8.3 | 5.7 |
| Agbogli et al. 2017<sup>49</sup> | Ashanti | Adults 18 years or above | 113 | WHO diagnostic criteria (fasting blood glucose only) | 3.5 | 5.9 | 2.5 | NR | NR |
| Chillunga et al. 2019<sup>39</sup> | Ashanti | Adults aged 25-70 years old | 1,436 | WHO diagnostic criteria | 5.7 | NR | NR | 8.8 | 3.6 |
| Opore-Addo et al. 2020<sup>44</sup> | Ashanti | Adults 18 years or above | 684 | Self-reported | 5.4 | NR | NR | NR | NR |
| Vuvor et al. 2011<sup>30</sup> | Greater Accra | Adults aged 36-95 years old | 597 | Urinary dipsticks and FBG determinations (level not mentioned) | 3.9 | 3.5 | 4.2 | NR | NR |
| Bawah et al. 2019<sup>26</sup> | Greater Accra | NA | 130 | HBA1C ≥ 6.5% | 5.4 | NR | NR | NR | NR |
| Quaicoe et al. 2017<sup>21</sup> | Volta | Adults aged 18-64+ years | 226 | American Diabetes Association Criteria 2010 | 8.6 | NR | NR | NR | NR |
| Bawah et al. 2019<sup>26</sup> | Volta | Adults 30 years or above | 202 | American Diabetes Association criteria 2010 | 6.9 | 6.5 | 7.1 | NR | NR |
| Ellahi et al. 2022<sup>29</sup> | Volta | Adults 18 and above | 850 | WHO diagnostic criteria | 4.4 | NR | NR | NR | NR |
| Gato et al. 2017<sup>33</sup> | Central | Adults 18-80 years | 482 | Self-reported | 8.3 | NR | NR | NR | NR |
| Anderson et al. 2017<sup>38</sup> | Central and Western | Adults 18 years or above | 976 | Fasting Blood Glucose ≥ 126mg/dl (7 mmol/L) | 24.6 to 39.8 | NR | NR | NR | NR |

NR- Not reported

#### 2) Common complications and comorbidity

Seven complications and/or comorbidities were identified in this review (figure 6). Studies on complications and comorbidities were only found for six of the ten administrative regions: Ashanti (n=9), Northern (n=2), Greater Accra (n=2), Volta (n=2), Brong-Ahafo (n=1) and Central (n=1). In addition, a multi-region (sub-national) study combining populations from Greater Accra, Ashanti, and Northern regions (n=1) was identified along with a study that did not specify location (n=1). No study was conducted at the national level for complications and comorbidities of diabetes.

**Figure 6:**
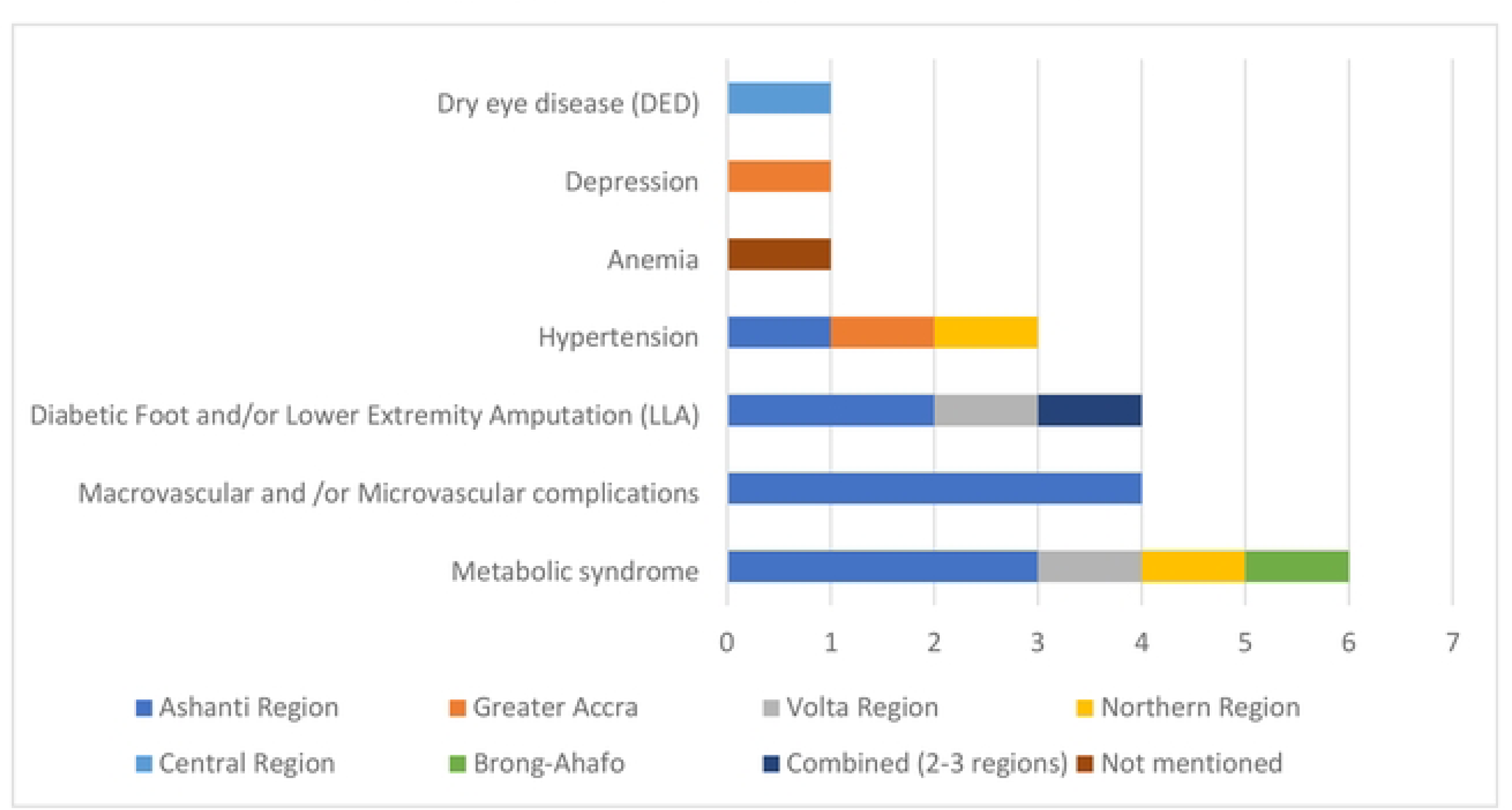
Number of studies per region by complication type.

The most common complication types reported in the included studies were metabolic syndrome (n=6), followed by macrovascular and/or microvascular complications (n=4) and diabetic foot and/or lower extremity amputation (n=4), hypertension (n=3), anaemia (n=1), depression (n=1) and dry eye disease (n=1). Figure 6 shows the number of studies per region by complication type.

The prevalence of micro and macrovascular complications among people with diabetes varied: coronary artery disease (CAD) ranged between 18.4%^37^ and 31.8%^41^, peripheral arterial disease (PAD) between 11.2%^37^ and 19%^17^, neuropathy between 18.3%^17^ and 20.8%^41^, nephropathy between 12.5%^41^ and 44.70%^17^, and retinopathy between 6.5%^41^ and 13.7%^45^.

Six studies assessed the prevalence of metabolic syndrome in diabetic patients in Ghana in the regions of Ashanti, Northern, Volta and Brong-Ahafo. The highest prevalence rate of metabolic syndrome was reported in the Ashanti region, with prevalence ranging between 42%^50^ and 90%^43^. This was followed by Brong-Ahafo with 68.6%^35^, the Volta region with 43.8%^27^, and Northern region with 24%^34^. The prevalence of hypertension among diabetic patients was assessed in three studies that included populations of Ashanti, Northern, and Greater Accra regions. The highest prevalence of hypertension as a complication/ comorbidity of diabetes was reported in the Greater Accra region (36.60%). This was followed by the Northern region (21%), and the Ashanti region (1.61%).

Diabetic foot disorders and lower extremity amputation were assessed in four studies. Two longitudinal studies were conducted in Komfo Anokye Teaching Hospital in the Ashanti region reporting a mean incidence of foot disorders and average incidence rate of diabetes related amputations of 8.39% (5.27% males and 3.12% females)^19^ and 2.4 (95% CI:1.84–5.61) per 1000 follow-up years^18^ respectively among diabetes patients. The third study was a cross-sectional study that randomly selected patients from the outpatient diabetes clinics of three tertiary hospitals from Greater Accra, Ashanti, and Northern regions and reported a prevalence of 11% for diabetic foot ulcers and 3% for lower extremity amputations. The fourth study was also cross sectional in Volta region focusing on diabetic foot ulcers^28^.

One study assessed the prevalence of depression among diabetic patients at the National Diabetes Management and Research Centre, Korle Bu Teaching Hospital in Greater Accra region^31^. The study reported that 31% of diabetic patients suffered from depression in 2018. Finally, a case-control study reported that 84.8 % of patients with diabetes had haemoglobin concentrations that were significantly lower than the general population. Table 3 provides a summary of findings stratified by complication type and geographic region in Ghana.

**Table 3:** Summary of studies that reported on the prevalence of complications/comorbidities for diabetics in Ghana.

| Metabolic syndrome |  |  |  |  |  |  |
| --- | --- | --- | --- | --- | --- | --- |
| Region | Author and date | Target population | Context | Sample size | Diagnostic criteria | Main results |
| Ashanti | Nsiah et al. 2015<sup>42</sup> | Adults aged 20-86 years old | T2D patients attending the Diabetic Centre of the Komfo Anokye Teaching Hospital in Kumasi, Ashanti region | 150 | NCEP/ATP III | The overall percentage prevalence of MetS was 58%. Males had a lower percentage prevalence of 22.99%, compared to a higher percentage prevalence of 77.01% for females. |
| Ashanti | Agyemang-Yeboah et al. 2019<sup>43</sup> | NA | Diabetic patients attending the Diabetic Clinic of the Komfo Anokye Teaching Hospital (KATH) Kumasi, Ashanti Region | 405 | NCEP/ATP III | The prevalence of metabolic syndrome observed among the study population was 90.6%. However, the MS condition among female participants (94.1%) was significantly higher than that of their male counterparts (76.5%) with $p < 0.0001$. |

| **Ashanti** | Odame Anto et al 2021<sup>50</sup> | Adults 30 years or above | Diabetic patients attending the Diabetic Clinic of the Komfo Anokye Teaching Hospital (KATH) Kumasi, Ashanti Region | 241 | NCEP/ATP III | The prevalence of metabolic syndrome observed among the study population was 42.7%. Among females, 52.8% (75/142) had MetS |
| --- | --- | --- | --- | --- | --- | --- |
| **Northern** | Mogres et al. 2014<sup>34</sup> | NA | Patients diagnosed with T2D receiving care from an outpatient clinic of the Tamale Teaching Hospital | 200 | IDF Consensus | The prevalence of MetS was 24.0% (n=48). The prevalence was higher in women (27.3%, n= 42) compared to men (13.0%, n=6). The commonest occurring components of the MetS included abdominal obesity (77.0%) and elevated FPG (77.0%) denoting uncontrolled diabetes. The prevalence of elevated BP was found to be 44.0%(n=88) and was higher in men (56.5%) than in women (40.3%). |
| **Volta** | Osei-Yeboah et al. 2017<sup>27</sup> | Adults aged 25-86 years old | Diabetic patients attending diabetic management clinic at the Ho Municipal Hospital in the Volta Region | 162 | NCEP-ATP III, the WHO, and the IDF criteria | The overall prevalence of metabolic syndrome among the study population was 43.83%, 63.58%, and 69.14% using the NCEP-ATP III, the WHO, and the IDF criteria, respectively. The most predominant component among the study population was high blood pressure using the NCEP-ATP III (108 (66.67%)) and WHO (102 (62.96)) criteria and abdominal obesity (112 (69.14%)) for IDF criteria. High blood pressure was the most prevalent component among the males while abdominal obesity was the principal component among the females. |
| **Brong-Ahafo** | Abagre et al<sup>35</sup> | Adults aged 30-79 years old | Diabetic patients enrolled at selected diabetes clinics (Dormaa Presbyterian and Berekum Holy Family Hospitals) | 430 | NCEP-ATP III, the WHO criteria | The prevalence of MS was 68.6% (95% CI: 64.0–72.8), higher among women (76.3%, 95% CI: 70.6–81.2) than men (58.0%, 95% CI: 35.0–49.4) and in the 50–59-year age group (32.1%) |
| **Diabetic foot and or lower extremity amputations** |  |  |  |  |  |  |
| Region | Author and date | Target population | Context | Sample size | Diagnostic criteria | Main results |

| **Ashanti** | Sarfo-Kantanka et al. 2018<sup>19</sup> | NA | Patients enrolled with the diabetes clinic of Komfo Anokye Teaching Hospital, a tertiary hospital in Kumasi. | 7,383 | Diabetic foot disorders include foot ulcers, PADs, and gangrene | The mean incidence of foot disorders was 8.39% (5.27% males and 3.12% females). An increase in the incidence of diabetic foot ranging from 3.25% in 2005 to 12.57% in 2016, $p < 0.001$, was determined. |
| --- | --- | --- | --- | --- | --- | --- |
| **Ashanti** | Sarfo-Kantanka et al. 2019<sup>18</sup> | NA | Patients enrolled with the diabetes clinic of Komfo Anokye Teaching Hospital, a tertiary hospital in Kumasi. | 3,143 | The global lower extremity amputation study | The average incidence rate of diabetes-related amputation was 2.4 (95% CI:1.84–5.61) per 1000 follow-up years: increasing from 0.6% (95% CI:0.21–2.21) per 1000 follow-up years in 2010 to 10.9% (95% CI:6.22–12.44) per 1000 follow-up years in 2015. |
| **Greater Accra, Ashanti, and Northern** | Atosona et al. 2019<sup>46</sup> | NA | Randomly selected patients from the outpatient diabetes clinics of three tertiary hospitals namely: Korle Bu Teaching Hospital, Komfo Anokye Teaching Hospital, and Tamale Teaching Hospital | 100 | International Consensus on Diabetic Foot | Among the patients, 11% had diabetic foot ulcers whilst 3% had lower extremity amputations. |
| **Volta** | Tuglo et al.<sup>28</sup> | NA | Diabetic patients attending selected diabetic clinics (Ho Teaching Hospital, Ho Municipal Hospital, Hohoe Municipal Hospital, and Margret Marquart Catholic Hospital) | 473 | Not mentioned | Foot ulcers were observed in 41 (8.7%) diabetic patients |
| **Hypertension** |  |  |  |  |  |  |
| **Region** | **Author and date** | **Target population** | **Context** | **Sample size** | **Diagnostic criteria** | **Main results** |
| **Ashanti** | Opere-Addo et al. 2020<sup>44</sup> | Adults 18 years or above | Rural districts in the Ashanti region of Ghana (Amansie West and Offinso North, Asante Akim South and Ahafo Ano South) | 684 | NA | The prevalence of hypertension was 111 (16.23%). Diabetes was prevalent in 37 (5.41%) of the study participants; thus, the prevalence of hypertension and diabetes was 137 (20.02%). The prevalence of diabetes and hypertension as a comorbidity was 11 (1.61%). |

| **Greater Accra** | Nimako et al. 2018<sup>32</sup> | Adults 18 years or above | Patients attending Tema General Hospital (TGH) in the Greater Accra Region | 1,527 | NA | The prevalence of multimorbidity was 38.8%, and around half (48.6%) of the patients with multimorbidity were aged between 18–59 years old. The most common combination of conditions was hypertension and diabetes mellitus (36.6%), hypertension and musculoskeletal conditions (19.9%), and hypertension and other cardiovascular conditions (11.4%). |
| --- | --- | --- | --- | --- | --- | --- |
| **Northern** | Mogre et al. 2014<sup>34</sup> | NA | Previously diagnosed diabetes mellitus patients attending a diabetic clinic at the Tamale Teaching Hospital | 100 |  | In general, 7.0% of the participants were underweight and 32.0% were overweight or obese. 21% of the studied participants were hypertensive. Prevalence of hyperglycaemia was higher among patients aged ≤40 years (88.9% vs. 75.8%) |
| **Micro and Macro vascular complications of DM** |  |  |  |  |  |  |
| **Region** | **Author and date** | **Target population** | **Context** | **Sample size** | **Diagnostic criteria** | **Main results** |
| **Ashanti** | Sarfo-Kantanka et al. 2016<sup>17</sup> | NA | Diabetes admissions at Komfo Anokye Teaching Hospital (KATH) in Kumasi | 11,414 | NA | Two thousand three hundred and ninety-two (21.0%) diabetic admissions were due to end-organ complications. Of these, 503 (18.7%) had peripheral vascular diseases, 377(14.0%) had coronary artery diseases, peripheral neuropathic ulcers (26.4%), 529 nephropathies (18.3%), 282 (10.5%) cerebrovascular diseases. Again 1207(44.8%) had nephropathy and 325(12.0) had peripheral neuropathic ulcers. |
| **Ashanti** | Lartey et al. 2018<sup>45</sup> | Adults 18 years or above | Diabetic Center and the Eye Department of a tertiary teaching hospital in the Ashanti region of Ghana | 208 | NA | Non-insulin-dependent diabetics constituted 97.1% whilst 2.9% were insulin-dependent diabetics. The prevalence of the outcome measures was: Cataract (23.7%) mild and moderate retinopathy (13.7%) severe proliferative retinopathy (1.8%) maculopathy (6.8%). The prevalence of low vision and blindness was 18.4%. Amongst diabetics, 59.1% had no previous eye evaluation. Impaired vision due to cataracts was 24.0 % |

|  |  |  |  |  |  | representing a 40% decline in a decade. |
| --- | --- | --- | --- | --- | --- | --- |
| **Ashanti** | Annani-Akollor et al. 2019<sup>41</sup> | Adults 18 years or above | Ghanian T2DM adults at Kmofo Anokye Teaching hospital (KATH) | 1,600 | NA | The prevalence of macrovascular and microvascular complications of T2DM was 31.8% and 35.3% respectively. The prevalence of neuropathy, nephropathy, retinopathy, sexual dysfunction, diabetic ketoacidosis (DKA), and hypoglycemia were 20.8%, 12.5%, 6.5%, 3.8%, 2.0%, and 0.8% respectively. The prevalence of single, double, and multiple complications are 59%, 16.3%, and 1.5% |
| **Ashanti** | Hayfron - Benjamin et al. 2019<sup>37</sup> | Adults aged 25-70 years old | Ghanaian adult T2DM population (206 in Ghana) aged >25 years | 650 | Nephropathy based on report from Joint Committee on Diabetic Nephropathy; PAD based on AHA 2012; coronary artery disease (CAD) was assessed using the WHO Rose angina questionnaire; Retinopathy, possible myocardial infarction, angina, and stroke were based on questionnaire | Microvascular and macrovascular complications rates were higher in non-migrant Ghanaians than in migrant Ghanaians (nephropathy 32.0% vs. 19.8%; PAD 11.2% vs. 3.4%; CAD 18.4% vs. 8.3%; and stroke 14.5% vs. 5.6%), except for self-reported retinopathy (11.0% vs. 21.6%) |
| **Other (Depression and Anaemia)** |  |  |  |  |  |  |
| Region | Author and date | Target population | Context | Sample size | Diagnostic criteria | Main results |

| **Greater Accra** | Akpalu et al. 2018<sup>31</sup> | Adults aged 30-65 years old | Patients recruited at the National Diabetes Management and Research Centre, Korle Bu Teaching Hospital, Accra, Ghana | 400 | Patient Health Questionnaire-9 (PHQ-9) | The prevalence of depression was 31.3% among T2DM patients. Female gender, being unmarried, frequent intake of alcohol, previous smoking status and insulin use were associated with increased odds of depression, whereas being educated above basic school level was associated with decreased odds of depression. |
| --- | --- | --- | --- | --- | --- | --- |
| **NA** | Antwi-Bafour et al. 2016<sup>20</sup> | NA | 50 control and 50 diabetic cases | 100 | NA | Of the patients with diabetes, 84.8 % had a haemoglobin concentration (incidence) that was significantly less (males 11.16±1.83 and females 10.41±1.49) than the controls (males 14.25±1.78 and females 12.53±1.14). |
| **Dry eye disease (DED)** |  |  |  |  |  |  |
| **Region** | **Author and date** | **Target population** | **Context** | **Sample size** | **Diagnostic criteria** | **Main results** |
| **Central** | Abu et al 2022<sup>51</sup> | Adults aged 38-85 years old | Diabetic patients attending Cape Coast Teaching Hospital | 311 | Diabetes was based on the American Diabetes association. DED Clinical assessment included meibum expressibility and quality, Schirmer test 1, tear breakup time (TBUT), ocular surface staining, and blink rates. | Prevalence of DED was 72.3% |

#### 3) Economic burden

Only one study reported on the economic burden of diabetes, assessing the financial cost of diabetes management (from a provider perspective) in cocoa clinics in Greater Accra, Ashanti, and Eastern regions^25^. Bottom-up micro costing was used to estimate the costs. The mean annual financial cost of managing one diabetic case at the outpatient clinics was estimated at GHS 540.3 (2021 US $194.09). The costs were broken down between service costs (22%) and direct medical costs (78%). Drug costs accounted for 71% of the direct medical costs. The cost of hospitalization per patient-day at clinics was estimated at GHS 32.78 (2021 US$ 11.78). The total annual financial cost of diabetes management accounted for 8% of the total annual expenditure of the clinics.

## Discussion

This scoping review reveals that there is paucity of literature on the burden of diabetes in Ghana. We divided the burden into three forms (themes): epidemiological burden (prevalence and incidence of diabetes); health outcomes (mortality, diabetes complications and comorbidity); and economic burden (cost of illness to the patients and health system). Most of the existing Ghana centred literature focuses on the prevalence of diabetes, including its complications and comorbidities. There is sparse data on the economic burden of the disease in Ghana. This review did not find any study that used generic health outcome measures such as DALYs or QALYs to estimate the diabetes burden. The existing literature is skewed towards a few particularly Ashanti, leaving some geographical regions of Ghana without any reporting (including, for example, Upper East, and Upper West regions).

Out of the identified studies, 36 studies focused on the prevalence of diabetes and associated complications; only one study reported economic-related findings, indicating a need for more costing studies, important also to support the development of economic evaluations. All diabetes studies were either on T2D or referred to diabetes generally without specifying the type. No studies explicitly targeting T1D were found. Our findings suggest important gaps in the Ghanaian scientific literature, and the need for further research to characterise the burden of diabetes in the country.

The national prevalence of diabetes in Ghana was reported in the studies to be between 2.80% and 3.95%^22–24^. The reported national prevalence of diabetes (2.8-% to 3.95%) is below the sub-Saharan Africa regional average of 4.5%^3^. However, studies that reported on prevalence sub-nationally provided substantially higher estimates; for example in Western, Ashanti, and Central regions, diabetes prevalence was reported to be 39.8%^38^, 25.2%^48^ 24.6%^38^ respectively. Despite Ghana having seemingly lower levels of prevalence of diabetes at a national level in comparison to the SSA average, there remains an urgent need to put in place interventions to address these regional differences, and stem any further anticipated rise in disease burden^3^. The within-country regional variations call for a more targeted approach when implementing diabetes interventions.

Diabetes in Ghana was found to be more prevalent among women compared to men^22–24,30^. This is in line with the recently reported prevalence in SSA by International Diabetes Ferderation^3^ and other studies^52,53^. Systematic reviews have found that women were more likely to have diabetes [odds ratio1.65 (95% CI 1.43, 1.91)], and less likely to have glycaemic control than men. It has been argued that relative differences in physical activity between men and women may be a factor ^52^. There was an urban-rural divide in the prevalence of diabetes in Ghana where urban areas were reported to have a higher prevalence compared to the rural areas^22–24,39,40^. This finding is consistent with other studies done in India (prevalence of 15.0% and 19.0% in rural and urban areas respectively in the year 2015-2019)^54^, and Myanmar (prevalence of 7.1% and 12.1% in rural and urban areas respectively in year 2013/2014)^55^. This has been attributed to differences in dietary habits and levels of physical activity between urban and rural areas.

Hypertension and metabolic syndrome were among the most prevalent comorbidities of diabetic patients in Ghana. Hypertension prevalence was highest in the Greater Accra region (36.60% of diabetic patients)^32^. Metabolic syndrome is a cluster of conditions that include combinations of hypertension, central obesity, insulin resistance, or atherogenic dyslipidaemia^56^. The two main risk factors of metabolic syndrome are the increase in consumption of high-calorie, low-fibre fast food and a decrease in physical activity which may be linked to mechanized transportation and a sedentary form of leisure time activities. These are the same established behavioural risk factors for diabetes and obesity that are typically predominant in urban areas. Without treatment, diabetes, high blood pressure, and obesity can damage blood vessels, leading to micro and macrovascular complications, which can occur concurrently. The highest prevalence of metabolic syndrome was reported in the Ashanti region where it was reported to range from 59 to 90% among diabetic patients^42,43^.

Our results also align well with the Institute of Health Metrics and Evaluation (IHME) assessment of the trend of disease burden in Ghana. Notably, in the year 2000, diabetes was not within the top 10 disease groups in terms of burden of disease. By 2019 diabetes had moved up to eighth position and was linked to more than 2,157 DALYs per 100,000 people. Cardiovascular diseases, which are often complications of diabetes and metabolic syndrome, were ranked first in 2019, causing an estimated 6,216 DALYs per 100,000 population^57^. This evidence points to the rapid rise of diabetes, cardiovascular diseases and other NCDs, linked with common genetic, metabolic, and behavioural risk factors. The ranking also suggests that there is a gradual receding of communicable diseases in the last 2 decades compared to NCDs. Combined, diabetes and cardiovascular diseases is linked to more than 8,300 DALYs per 100,000 people in Ghana, representing a significant proportion (12.6%) of the total disease burden in the country^57^.

Literature on the economic burden of diabetes in Ghana is very limited with only one study reporting on the burden. This finding is in line with the study by Hollingworth et al. (2020) that looked at available localised information to support HTA in Ghana noting that there were few accessible data sources for costs and resource utilisation generally^9^. Relatedly, we found a falling off in the number of studies published after 2019, although that may reflect the impact of the COVID-19 pandemic on research and publication choices within the country and globally

### Policy implications

Our review provides some evidence of the situation in Ghana, and associated information gaps, consistent with studies focused on other African countries^58^. We find that Ghana is faced with a rising prevalence of diabetes and cardiovascular disorders with potentially important regional differences in disease burden. To address this challenge, there is a need to understand the contextual factors driving the rise and the likely causes behind the existing regional variations in the reported burden.

Unlike communicable diseases, NCDs are usually chronic in nature and exhibit a progressive disease course. A person may develop more than one NCD at a time, fuelling disease progression even further^59,60^. NCDs constitute a long-term burden not only to the patient and carers, but also to the healthcare system and the economy. Currently, NCDs do not attract as much development assistance funds as communicable diseases such as HIV, tuberculosis and malaria, and Ghana which is currently classed as a middle-income country is no longer eligible for some development assistance grants. Therefore, there is a need for Ghana, as well as countries, to ensure that available domestic resources for health can be used as efficiently as possible to address this growing burden. Typically, this involves operationalising and institutionalising proven priority-setting processes.

HTA can be applied to both treatments for NCDs, such as insulin analogues for the management of diabetes, and also to interventions that seek to reduce disease prevalence and incidence in the first place. On the latter this could include identifying cost-effective interventions to tackle the common NCD risk factors shared by diabetes and other comorbidities such as cardiovascular diseases, obesity, or metabolic syndrome to reduce the burden of diabetes in the country. This may require stronger prevention approaches^61^ targeting high risk individuals or whole populations, which aim to increase physical activity and promote a healthy diet while also monitoring obesity levels. This would be especially important in urban communities in Ghana where the burden appears higher than the national average.

Institutionalising HTA for localised decision making requires that relevant data sources are available, and that may mean developing a strategy to address key informational gaps as part of building any HTA system. There was a scarcity of evidence on the economic burden of diabetes in Ghana, and there were no estimates available of the burden of disease for a number of regions. A key issue is the lack of diabetes incidence data: as previously reported^9^, the Ghana Health and Demographic Surveillance Systems (GHDSS) could potentially be a valuable source of such information, and the Ministry of Health and other stakeholders could consider enhancing their operation in this space, for diabetes as well as other NCDs. It may also mean better leveraging already existing data sources, such as from the National Health Insurance Scheme^9^.

### Limitations

The scoping review did not include an appraisal of the quality of included studies. According to Arksey and O’Malley’s (2005) framework, the study quality is not assessed during scoping reviews, but rather in systematic reviews that aim to address specific questions relating to feasibility, appropriateness, meaningfulness or effectiveness of a certain treatment or practice. The exploratory nature of our study about the burden of diabetes made the scoping review methodology suitable.

The included studies used different diagnostic criteria for diabetes including WHI criteria, American Diabetes Association and self-reporting. Self-reported diabetes can be misleading because some of the people involved in the studies may not have received a diagnosis and thus report as not having the diabetes. This may have underestimated the prevalence of diabetes in the country or region.

This study has used an older geographical classification system for the administrative regions in Ghana. The sub-national regions of Ghana constitute the first level of subnational government administration within the country. From 1987, Ghana had ten officially established regional boundaries. In 2018, a referendum on the creation of six new regions was held and the overall number were later increased to 16. We used the older system of 10 regions based on the available literature which mostly reported within that classification system. However, we expect that there would be no substantial differences had the newer system been applied, since all new regional divisions stem from the partitioning of regions where no studies were found (e.g, Brong Ahafo and Northern regions).

## Conclusion

Ghana is faced with a considerable burden of diabetes which varies by geographical region and setting (rural/urban). It is urgent to tackle the growing challenge to mitigate the likely enormous burden and cost of the disease. Despite the existing regional variation of the burden of diabetes, there is a paucity of literature in some regions (for example Eastern, Western, Upper East, Upper West, and Brong Ahafo). There is therefore a need for further research to understand the burden (epidemiological, health outcomes and economic) of diabetes in these regions in order to inform the NCD prevention and management policies in the country.

## Declaration

## Ethics approval and consent to participate

The study utilised secondary data/literature that is publicly available and did not use any personal or private data.

## Consent for publication

Not applicable

## Availability of data and materials

The extracted data analysed during the current study is available from the corresponding author upon reasonable request.

## Authors contributions

Conceptualisation: – MG, JK, FR

Developing and carrying out search strategy: – MG

Screening of articles: – MG, JK

Data extraction, and quality assessment: – MG, JK

Validation: – STR

Data curation and analysis: – MG, JK

Funding acquisition: – FR

Methodology: – MG, JK,

Project administration: – MG, JK, IA, GG, RO, EAK

Supervision: – FR, AV

Writing – original draft: – MG, JK,

writing review & editing: – MG, JK, RO, EAK, GG, IA, FR, STR, AV

## Data Availability

Data is available upon request

## Acknowledgements

None

## Annex

### Annex 1 Search strings

#### Web of Science search

Search link for Web of Science https://www.webofscience.com/wos/woscc/summary/55ba5b67-9492-4899-a3dd-8b2fe3588006-80f3249d/relevance/1

(TS=(Ghana)) AND TS=(Diabetes)

#### PubMed search string

((“ghana”[MeSH Terms] OR “ghana”[All Fields] OR “ghana s”[All Fields]) AND (“diabete”[All Fields] OR “diabetes mellitus”[MeSH Terms] OR (“diabetes”[All Fields] AND “mellitus”[All Fields]) OR “diabetes mellitus”[All Fields] OR “diabetes”[All Fields] OR “diabetes insipidus”[MeSH Terms] OR (“diabetes”[All Fields] AND “insipidus”[All Fields]) OR “diabetes insipidus”[All Fields] OR “diabetic”[All Fields] OR “diabetics”[All Fields] OR “diabets”[All Fields])) AND (2021/4/4:2023/4/11[pdat])

#### Embase search string

(‘ghana’/exp OR ghana) AND (‘diabetes mellitus’/exp OR ‘diabetes mellitus’)

#### Scopus search string

(TITLE-ABS-KEY ( ghana ) AND TITLE-ABS-KEY ( diabetes ) ) AND PUBYEAR > 2020 AND PUBYEAR < 2024

